# Cross-ancestry meta-analysis of opioid use disorder uncovers novel loci with predominant effects on brain

**DOI:** 10.1101/2021.12.13.21267480

**Authors:** Rachel L. Kember, Rachel Vickers-Smith, Heng Xu, Sylvanus Toikumo, Maria Niarchou, Hang Zhou, Emily Hartwell, Richard C. Crist, Christopher T. Rentsch, VA Million Veteran Program, Lea K. Davis, Amy C. Justice, Sandra Sanchez-Roige, Kyle M. Kampman, Joel Gelernter, Henry R. Kranzler

## Abstract

Despite an estimated twin heritability of ∼50%, genome-wide association studies (GWAS) of opioid use disorder (OUD) have revealed few genome-wide significant (GWS) loci, with replicated findings only in European-ancestry individuals. To identify novel loci, including those in non-European ancestries, and improve our understanding of the biology of OUD, we conducted a cross-ancestry meta-analysis using the Million Veteran Program (MVP). OUD cases in MVP had at least 1 International Classification of Diseases (ICD)-9 or ICD-10 code for opioid abuse or dependence (N=31,473). Opioid-exposed controls (N=394,471) had one or more outpatient opioid prescription fills. We conducted GWAS for each major ancestral group in MVP: African Americans (AAs; N=88,498), European Americans (EAs; N=302,585), and Hispanic Americans (HAs; N=34,861), followed by a cross-ancestry meta-analysis. Ten loci were GWS in the cross-ancestry meta-analysis, 8 of them novel. In addition to the known coding variant rs1799971 in *OPRM1*, which was the lead SNP genome-wide (p=6.78×10^−10^), and a recently reported exonic variant in *FURIN*, we identified intronic variants in *RABEPK, FBXW4*, *NCAM1*, and *KCNN1*. Ancestry-specific analyses identified an additional novel locus for each of the 3 ancestry groups. A supplementary meta-analysis within EAs that included MVP and other samples identified a locus in *TSNARE1*, which was also GWS in the cross-ancestry meta-analysis of all datasets. Gene-based association analyses identified 1 gene in AAs (*CHRM2*) and 3 in EAs (*OPRM1, DRD2*, and *FTO)*. Significant genetic correlations (*r*_g_’s) were identified for 127 traits, including positive correlations with schizophrenia, problematic alcohol use, and major depressive disorder. The most significantly enriched cell type group was the central nervous system with gene-expression enrichment identified in brain regions previously associated with substance use disorders. With a case sample 50% larger than that of the previous largest GWAS, we identified 14 loci for OUD, including 12 novel loci, some of which were ancestry-specific. These findings increase our understanding of the biological pathways involved in OUD, which can inform preventive, diagnostic, and therapeutic efforts and thereby help to address the opioid epidemic.

## Introduction

Opioid use disorder (OUD) is a problematic pattern of opioid use that leads to significant impairment or distress^1^. In the United States, a 10-fold increase in opioid analgesic prescriptions between 1990 and 2010 contributed to an epidemic of opioid misuse, abuse, and overdose deaths^2–4^. By 2019, 3.7% of U.S. adults reported past-year opioid misuse and 0.6% met criteria for an OUD^5^. Overdose deaths, which continue to increase annually, have reached crisis proportions^6^, reflecting the limitations of available preventive and treatment efforts.

Genetic studies can help inform our understanding of the biology underlying OUD. However, although the estimated heritability (h^2^) of OUD based on twin and family studies is ∼50%^7^, few genetic associations have been identified. Genome-wide association studies (GWAS) of OUD, opioid dependence (OD) or related phenotypes have yielded inconsistent results, likely due to the limited sample size of the discovery datasets and different case and control definitions^8–11^.

The use of data from electronic health records (EHRs) linked to biobanks has permitted research consortia to assemble increasingly large GWAS samples. An EHR-based study of 1,039 OUD cases and 20,271 controls identified two genome-wide significant (GWS) loci, with common SNPs explaining 6.0% of the phenotypic variation in OUD^12^. A meta-analysis of European Americans (EAs) (10,544 OUD cases and 72,163 opioid-exposed controls) and African Americans (AAs) (5,212 OUD cases and 26,876 opioid-exposed controls) based largely on data from the Million Veteran Program (MVP), identified a single GWS SNP, rs1799971 in *OPRM1*, in EAs only, with SNP-based heritability estimated at 11.3%^13^. There were no GWS findings in AAs or in a cross-ancestry meta-analysis. A study that combined data from multiple cohorts (20,858 OUD cases), including an earlier release of MVP data, identified 2 GWS loci – a variant within *OPRM1* in a cross-ancestry analysis, and an additional variant in *FURIN* in a European-ancestry meta-analysis^14^.

Two recent GWAS have increased sample sizes for genetic discovery by examining opioid-related phenotypes other than OUD. A GWAS of prescription opioid misuse in an EA sample from 23andMe (27,805 cases) identified 2 novel GWS loci^15^. A meta-analysis of European-ancestry individuals included 23,367 cases ascertained using either Diagnostic and Statistical Manual of Mental Disorders diagnoses (DSM) or frequency of opioid use (FOU) and 384,629 controls^16^. Using a latent trait [opioid addiction (OA)] generated from a genomic structural equation model to conduct GWAS, the group identified GWS SNPs in *OPRM1* and, in gene-based analyses, *PPP6C* and *FURIN*. The SNP h^2^ for the DSM trait was 11% and 18% for the FOU trait.

These studies also identified significant genetic correlations (r_g_’s) with traits well known to co-occur with OUD, suggesting widespread pleiotropy. The strongest positive r_g_’s were with substance-related traits (e.g., alcohol dependence/use disorder, cannabis use disorder, drinks/week, cigarettes/day)^12, 13, 15, 16^, and psychiatric disorders (e.g., attention deficit hyperactivity disorder (ADHD), posttraumatic stress disorder (PTSD), major depressive disorder (MDD), schizophrenia, bipolar disorder, neuroticism)^13, 15, 16^. Although these traits are known to be genetically correlated, it is unknown whether there is a shared genetic structure between OUD, other substance use, and psychiatric disorders. Negative r_g_’s were seen for educational attainment^13, 15, 16^, cognitive performance^13^ and subjective wellbeing^16^. Causal effects on OUD for some of these traits were identified via Mendelian randomization (MR) analysis. Positive causal effects on OUD were found for the following exposures: regular tobacco smoking, depressed affect, neuroticism, and cognitive performance. A negative causal effect on OUD was also seen with cognitive performance as the exposure^13^. The causal effect of OUD on these traits was unable to be examined due to the limited number of GWS variants.

The different phenotypes used in these studies reflect the difficulty of ascertaining a large, multi-ancestry, well-characterized sample for use in GWAS of opioid-related phenotypes. EHR-based traits generally use International Classification of Disease (ICD) diagnostic codes for phenotyping OUD (e.g.,^12, 13^). Cohorts recruited from some non-clinical biobanks rely on single-item, self-report questionnaires (e.g.,^15^) or have combined multiple case and control definitions derived as latent variables in genomic structural equation modeling^16^. A key consideration in selecting OUD cases, particularly given the high prevalence of opioid use in the United States, is the stringency of the definition. More stringent case definitions increase confidence in the specificity of the diagnosis and, by reducing heterogeneity, may increase statistical power. However, they also reduce the sample size and have the potential to reduce generalizability by not capturing a disorder’s full range of presentations (e.g., by misclassifying cases as subthreshold).

Here, we conducted a cross-ancestry meta-analysis of OUD that included AA, EA, and Hispanic American (HA) subjects recruited from the MVP that maximized OUD cases by using a less stringent definition (requiring the presence of a single OUD diagnostic code) and compared them to opioid-exposed controls (N cases=31,473, N controls=394,471). In supplementary analyses, we compared our results to those using a stringent OUD phenotype in MVP, and performed a meta-analysis that combined data from the MVP, Yale-Penn (unpublished data), the Partners HealthCare Biobank^12^, and the Psychiatric Genomics Consortium (PGC)^11^. To elucidate the biology implicated by the variation that we identified through GWAS, we performed gene-based, gene-set enrichment and transcriptome-wide association analyses; examined r_g_s with a wide variety of phenotypes; calculated polygenic risk scores (PRS) and performed phenome-wide association studies (PheWAS) in independent samples; and conducted Mendelian randomization analyses and genomic structural equation modeling to evaluate the causal effect and shared genetic structure between OUD, substance-related traits, and psychiatric disorders. We identified GWS findings in AA, EA and HA individuals. Further, we obtained novel insights that implicate genes expressed in the central nervous system (CNS), including specific brain regions implicated in the biology of OUD.

## Methods

### Overview of Analyses

We conducted an ancestry-specific GWAS using a less stringent OUD case definition in AAs, EAs, and HAs from the MVP, followed by a cross-ancestry meta-analysis. Cases had received at least one lifetime ICD Ninth Revision (ICD-9) or Tenth Revision (ICD-10) diagnosis of OUD and control subjects were opioid exposed. Further details on phenotyping are described below. This GWAS was used for all subsequent downstream analyses.

In a supplementary analysis, we performed within-ancestry meta-analyses for AAs and EAs from the MVP, Yale-Penn (unpublished data), the Partners HealthCare Biobank^12^, and the PGC^11^, followed by a cross-ancestry meta-analysis which included all samples. In a second supplementary analysis, we repeated the GWAS in MVP with the more stringent case definition used in the prior MVP OUD GWAS^13^. An overview of the analyses is provided in Supplementary Figure 1.

### Million Veteran Program Cohort

As of September 2021, the MVP^17^ had recruited approximately 850,000 veterans at 63 VA medical centers nationwide. All participants provide written informed consent and a blood sample for DNA extraction and genotyping and give permission to access their EHR for research purposes. The MVP was approved by the Central Veterans Affairs Institutional Review Board (IRB) and all site-specific IRBs. All relevant ethical regulations for work with human subjects were followed in the conduct of the study.

### Phenotypes

OUD diagnostic codes based on ICD-9/10 were obtained from the VA EHR. The main GWAS used a less stringent definition of OUD (N = 31,473), which required the presence of 1 inpatient or outpatient ICD-9/10 diagnostic code for OUD (304.0, 304.7, 305.5, F11.1, F11.2) in the EHR. The stringent definition (N = 23,459), used in the supplementary GWAS, required at least 1 inpatient or 2 outpatient ICD-9/10 OUD diagnostic codes in the EHR. Controls (N = 394,471) for all GWAS were defined as individuals with at least 1 outpatient opioid analgesic prescription fill [excluding prescriptions for OUD treatment (e.g., buprenorphine or methadone)] and no ICD-9/10 diagnosis code for OUD documented in the EHR (i.e., opioid-exposed). In all analyses, age at the time of MVP enrollment was used as a covariate. Demographics are presented in Supplementary Table 1.

### Genotyping and Imputation

The genotyping of samples in the MVP is ongoing and, in this analysis, we used Release 4 imputed data. MVP samples were genotyped with a custom Affymetrix Axiom Biobank Array. Quality control of genotype data and subsequent imputation were performed by the MVP Genomics working group. Duplicate samples were removed, as were those with a sex mismatch, 7 or more relatives in MVP, excessive heterozygosity or a genotype call rate <98.5%. Variants were removed if they were monomorphic, had a missing call rate <0.8, or a Hardy-Weinberg equilibrium p<1×10^−6^ both in the entire sample using a PCA-adjusted method and within 1 of the 3 major ancestry groups (AA, EA, HA). Genotypes were phased with SHAPEIT4 (v.4.1.3)^18^ and imputed using Minimac4^19^, with biallelic SNPs imputed using the African Genome Resources (AGR) reference panel by the Sanger Institute, and non-biallelic SNPs and indels imputed in a secondary imputation using the 1000 Genomes Project phase 3, version 5^20^ reference panel. Indels and complex variants from the second imputation were merged into the AGR imputation.

Genome-wide association analyses were performed using PLINK 2.0^21^. We removed 1 individual from each pair of related individuals at random (kinship>0.08, N=31,010). Genetic ancestry was unified with self-identified race/ethnicity using the HARE (Harmonizing Genetic Ancestry and Self-Identified Race/Ethnicity) method^22^. Quality control of imputed variants was performed within each ancestral group. Genetic variants were excluded based on minor allele frequency (MAF: AA<0.005; EA<0.001; HA<0.01), genotype call rate<0.95, and Hardy-Weinberg equilibrium p<1×10^−6^ or a population-specific imputation quality (INFO) score <0.7. Covariates included sex, age at enrollment, and the first 10 genetic principal components (PCs) within each ancestry.

### Datasets for Meta-analysis

Supplementary Table 2 summarizes the datasets used for meta-analysis. Summary statistics for GWAS of OUD were obtained from two previously published datasets: 1) Partners HealthCare Biobank, which used the same less stringent case definition and opioid-exposed controls in European ancestry individuals^12^ 2) PGC, which used a DSM-IV OD diagnosis and opioid-exposed controls in African and European ancestry individuals^11^. We also included the Yale-Penn 3 (YP3) unpublished dataset (Yale-Penn 1 and 2 were included in PGC analyses). In YP3, we conducted a GWAS using cases with a DSM-IV OD diagnosis and opioid-exposed controls. For AAs, there were 168 cases and 153 controls; for EAs, there were 578 cases and 219 controls. We used GEMMA to conduct association analysis to account for relatedness between the individuals. Sex, age at recruitment, and the first 10 PCs were included as covariates.

### Meta-analysis and Independent Variants

Meta-analyses were conducted using a sample-size-weighted method in METAL^23^. To compensate for the imbalance in the ratio of cases to controls, effective sample sizes were calculated using the formula: 4/[1/n_case + 1/n_control]. Effective sample sizes were used in all meta-analyses and all downstream analyses. Meta-analyses were conducted across the following datasets: 1) cross-ancestry (AA, EA, and HA) meta-analysis within MVP, 2) within-ancestry meta-analysis across datasets (AA: MVP, PGC, YP3; EA: MVP, Partners HealthCare Biobank, PGC, YP3), 3) cross-ancestry meta-analysis across all datasets (AA [MVP, PGC, YP3]; EA [MVP, Partners HealthCare Biobank, PGC, YP3]; HA [MVP]).

To identify independent variants, we performed LD-clumping within each ancestry using a range of 3000 kb, r^2^ > 0.1, and the matched 1000 Genomes^20^ reference panel. Following clumping, variants that were located <1Mb apart were merged into a single locus. For loci that contained multiple variants, we conducted conditional analyses using COJO in GCTA^24^. Within each locus, we conditioned on the most significant variant. Upon conditioning, variants within the locus that remained significant (p < 5 × 10^−8^) were considered independent.

### SNP-based Heritability Analyses and Partitioning Heritability Enrichment

We used LD score regression^25^ (LDSC) to estimate OUD SNP-based heritability (h^2^_SNP_) in AAs and EAs for common SNPs mapped to HapMap3^26^. To ensure matching of population linkage disequilibrium (LD) structure, we used pre-computed LD scores based on African and European 1000 Genomes Project Phase 3^20^. SNPs in the major histocompatibility complex (MHC) region were excluded. Because of the high degree of genetic admixture in HAs and the smaller size of the sample, we did not estimate h^2^_SNP_ in that population group.

We used LDSC to partition h^2^_SNP_ in the OUD EA dataset and examined the enrichment of the partitioned h^2^_SNP_ based on different functional genomic annotation models^27, 28^. In the baseline model, we examined 53 overlapping functional annotations comprising genomic, epigenomic, and regulatory features. Next, we analyzed 10 overlapping cell-type groups derived from 220 cell-type-specific annotations. Finally, enriched cell-type categories were analyzed based on annotations obtained from H3K4me1 imputed, gapped peak data generated by the Roadmap Epigenomics Mapping Consortium^29^. For each h^2^_SNP_ partitioning model, multi-allelic and MHC region variants were excluded, and Bonferroni-correction was applied to identify significant enrichment.

### Gene-based, Functional Enrichment and Pathway Analyses

We performed gene-based association testing for OUD in FUMA v1.3.6a^30^, using MAGMA v1.08^31^, which employs multiple regression models to detect multiple marker effects that account for SNP p-values and LD between markers, using the matched-ancestry 1000 Genomes Project phase 3^20^ panel as LD reference. We used a total of 18,707 protein-coding genes, with p < 2.67 × 10^−6^ (0.05/18,707) considered GWS.

To identify gene sets enriched for OUD, we used MAGMA^31^ to curate gene sets; Gene Ontology terms (obtained from MsigDB c2); and GWAS-catalog enrichment, correcting for gene size, variant density, and LD within and between genes. We also used MAGMA to test the association between gene-set properties and tissue-specific gene expression profiles using GTEx (v.7) data from 53 tissues (Bonferroni-corrected p-value threshold = 9.43 x 10^−4^).

### Transcriptome-wide Association Analyses

We performed transcriptome-wide association analyses using the MetaXcan framework^32^ and the GTEx release v.8 eQTL MASHR-M models^33^. Forty-nine tissues from GTEx v.8 were analyzed comprising 12,951 samples. First, GWAS summary statistics were harmonized for the EA population based on the human genome assembly GRCh38 (hg38) and linked to the 1000 Genomes reference panel using GWAS tools (https://github.com/hakyimlab/summary-gwas-imputation/wiki/GWAS-Harmonization-And-Imputation). A transcriptome-wide association analysis of 49 tissues was run using S-PrediXcan^32^. A Bonferroni correction for statistical significance was applied within each tissue conditioned on the number of genes tested (Supplementary Table 14).

Because expression quantitative trait loci (eQTL) were correlated across tissues, we integrated gene expression signals for 49 tissue panels using S-MultiXcan^34^ and tested 10,552 genes in total. Resulting p-values were Bonferroni corrected to identify significant gene associations (p-value threshold = 4.74 × 10^−6^).

### Drug Interactions

To identify drugs that could potentially be repurposed to treat OUD, we examined genes identified in the variant or gene-level analyses using the Drug Gene Interaction Database^35^ (https://www.dgidb.org). Medications were categorized using the Anatomical Therapeutic Chemical (ATC) classification system, retrieved from the Kyoto Encyclopedia of Genes and Genomics Kyoto Encyclopedia of Genes and Genomics (KEGG; https://www.genome.jp/kegg/drug/).

### Genetic Correlation

We used LD score regression^25^ to calculate the *r*_g_ between a) OUD or OD datasets used for meta-analysis (AA [MVP, PGC, YP3]; EA [MVP, Partners HealthCare Biobank, PGC, YP3]) and b) OUD (MVP EA) and 40 other published traits comprising psychiatric, substance use, cognitive, and anthropometric traits selected based on a priori hypotheses (See Supplementary Table 20 for a full list), using pre-computed LD scores for HapMap3^26^ SNPs based on the matched-ancestry 1000 Genomes Project Phase 3^20^ reference panel. To explore additional traits in a hypothesis-free manner, we also estimated the *r*_g_ between OUD and 1,270 traits (comprising published and unpublished traits from the UK Biobank (UKBB) using the Complex-Trait Genetics Virtual Lab (CTG-VL) (https://genoma.io). CTG-VL integrates publicly available GWAS summary statistics and utilizes the LDSC framework to calculate *r*_g_ between complex traits and diseases of interest^36^. A Bonferroni correction was applied within each LDSC and CTG-VL analysis, and traits with a corrected p-value < 0.05 were regarded as significantly correlated.

### Mendelian Randomization

We performed Mendelian randomization (MR) analysis using the *MendelianRandomization* package in R. Causal relationships between OUD and other traits were tested bidirectionally using three methods: Weighted Median, Inverse Variance Weighted and MR-Egger. We tested for pleiotropy using the MR-Egger intercept test. Instrumental variants were those associated with the exposure at p < 1 × 10^−5^. When the instrumental variants were not present in the outcome data, we identified the best-proxy variant (LD > 0.8). Variants with MAF < 0.01 or with no proxy with LD > 0.8 within 200 kb were removed. Each trait included more than 20 instrumental variables, which provides a robust estimate of causal effects. We considered causal effects as those for which at least 2 MR tests were significant after Bonferroni correction and that showed no evidence of violation of the horizontal pleiotropy test (MR-Egger intercept p > 0.05).

### Polygenic Risk Scores and Phenome-wide Association Studies

We calculated PRS for OUD in two independent datasets (Yale-Penn and BioVU) using PRS-continuous shrinkage (PRS-CS)^37^, followed by phenome-wide association analyses (PheWAS). In each dataset, OUD summary statistics from the matched ancestry were used to calculate PRS. Details for the analysis in each dataset are below.

#### Yale-Penn

We removed SNPs with INFO score < 0.7, MAF < 0.01 genotype call rate < 0.95, or an allele frequency difference between genotyping batches > 0.4, which left a total of 8,811,422 SNPs. We removed one individual from each pair of related individuals with pi-hat > 0.25. To estimate genetic ancestry, we calculated PCs on common SNPs between Yale-Penn and 1000 Genomes Project Phase 3^20^ using PLINK 1.9^21^. Subjects were assigned to an ancestry based on the distance of 10 PCs from the 1000 Genomes reference populations. The resulting data set included 4,922 AAs and 5,709 EAs. We excluded binary phenotypes with fewer than either 100 cases or 100 controls, and continuous phenotypes with fewer than 100 individuals. We conducted PheWAS by fitting logistic regression models for binary traits and linear regression models for continuous traits. We used sex, age at recruitment, and the top 10 genetic PCs as covariates. We applied a Bonferroni correction to control for multiple comparisons.

#### BioVU

We used de-identified clinical data from Vanderbilt University Medical Center’s (VUMC) Biobank (BioVU). Details on the quality control process have been described elsewhere^38^. The genotyping information that we used was from the Illumina MEG^EX^ array. Genotypes were filtered for SNP and individual call rates, sex discrepancies, and excessive heterozygosity using PLINK v1.9^21^. Imputation of the autosomes was conducted using the Michigan Imputation Server^19^ based on the Haplotype Reference Consortium reference panel. PCA using FlashPCA2 combined with CEU, YRI and CHB reference sets from 1000 Genomes Project Phase 3^20^ was conducted to determine participants of African and European Ancestry. The sample was then filtered for cryptic relatedness by removing one individual of each pair for which pi-hat>0.2. This resulted in 12,384 individuals of African ancestry and 66,903 individuals of European ancestry samples for analysis. We conducted PheWAS by fitting a logistic regression for each of the 1,335 disease phenotypes available in BioVU to estimate the odds of a diagnosis of that phenotype given the OUD PRS. Each disease phenotype (commonly referred to as “phecode”; https://phewascatalog.org/phecodes, Phecode Map 1.2) was classified using ICD 9 and 10 diagnostic codes to establish “case” status. For an individual to be considered a case, they were required to have two separate ICD codes for the index phenotype, and each phenotype needed at least 100 cases to be included in the analysis. The covariates included in the analyses were sex, median age of the longitudinal EHR measurements, and the top 10 genetic PCs. The project was approved by the VUMC Institutional Review Board (IRB #s 160302, 172020, 190418).

### Genomic Structural Equation Modeling

We performed Genomic Structural Equation Modeling^39^ for OUD, 3 other substance use traits (problematic alcohol use, cannabis use disorder, ever smoked regularly), and 7 psychiatric disorders (schizophrenia, bipolar disorder, major depressive disorder, autism spectrum disorder, attention deficit hyperactivity disorder, Tourette’s syndrome, and obsessive-compulsive disorder). We calculated a genetic covariance matrix using multivariable LDSC and the 1000 Genomes Project phase 3 European samples^20^ as reference. An exploratory factor analysis was conducted using the genetic covariance matrix and a four latent-factor structure with varimax rotation. We used the determined structure containing paths with loading factor >0.2 to perform a confirmatory factor analysis implemented in the *GenomicsSEM* package in R. To prevent negative residual variance after estimation, we restricted the residual variance of OCD and ADHD to be greater than 0.

## Results

### Sample Description

Our MVP sample comprised 425,944 individuals (AA: 88,498; EA: 302,585; HA: 34,861), of whom 90.6% were male (Supplementary Table 1). The less stringent OUD definition yielded 28.8%-38.9% more cases across the ancestral groups (AA = 8,968, EA = 19,978, HA = 2,527) than the stringent definition (AA = 6,457; EA = 15,040; HA = 1,962). On average, less stringent cases had 77.2 (SD=96.9) opioid prescription fills, stringent cases had 76.5 (SD=97.6) fills, and controls had 25.0 fills (SD=48.3).

### Identification of Novel Loci Associated with Opioid Use Disorder

The cross-ancestry meta-analysis of the less stringent OUD diagnosis within the MVP sample yielded 12 GWS variants, 10 of which were independent after conditioning on the lead variant within each locus (Figure 1, Supplementary Table 3). The protein-coding genes nearest these variants are *CDKAL1*, *BTNL2*, and *OPRM1* (all on chr. 6), *RABEPK* (chr. 9), *FBXW4* and *7SK* (chr. 10), *NCAM1* (chr. 11), *FURIN* (chr. 15), *KCNN1* (chr. 19), and *RNF114* (chr. 20). The most robust signal was in *OPRM1* (lead SNP rs1799971, p=6.78 x 10^−10^), which replicates the main finding of the previous MVP OUD GWAS^13^. The variant in *FURIN* is supported by prior findings at the variant^14^ and gene-based^16^ levels. In addition, there were 3 ancestry-specific loci (Supplementary Table 4): 1 each in AAs (*NNT*, chr. 5), EAs (*CDH8*, chr. 16), and HAs (*MRS2*, chr. 8). 3 loci were GWS in the cross-ancestry meta-analysis of the stringent OUD diagnosis in the MVP sample, including 1 additional locus (*TSNARE1*, chr. 8) which was also found in the EA-specific analysis of the stringent OUD diagnosis (Supplementary Tables 5 and 6), and the meta-analysis of EA subjects’ data from MVP, Partners HealthCare Biobank, PGC and YP3 (Supplementary Table 8). GWS loci from all analyses are presented in Supplementary Tables 3-8. Based on all analyses, we identified a total of 14 GWS loci, 12 novel (Table 1).

**Figure 1:**
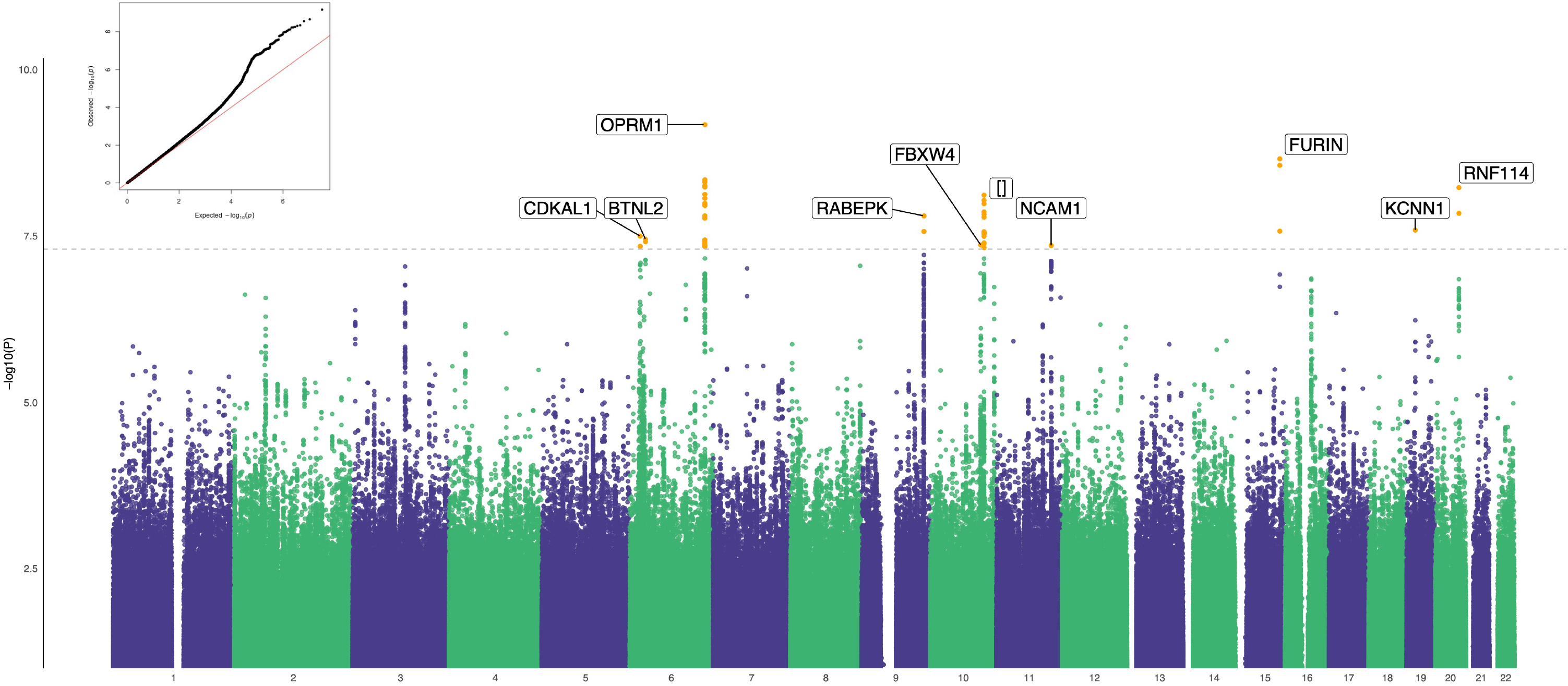
Manhattan and quantile-quantile plot for cross-ancestry meta-analysis of OUD (N case=31,480, N control=394,484). Effective sample size weighted meta-analyses were performed in METAL. The nearest protein-coding gene (<1Mb) in each locus is labelled.

**Table 1:** Summary of the 14 GWS loci identified in GWAS analyses of OUD

| Table 1: Summary of the 14 GWS loci identified in GWAS analyses of OUD |  |  |  |
| --- | --- | --- | --- |
| Chr | Position (GRCh37/hg19) of lead SNPs | Nearest genes | GWAS analysis |
| 5 | 43846681 | *NNT* | AA (Less stringent) |
| 6 | 21362610, 21478361 | *CDKAL1, SOX4* | Cross-ancestry (Less stringent), HA (Less stringent), Cross-ancestry (Meta) |
| 6 | 32383573 | *BTNL2* | Cross-ancestry (Less stringent) |
| 6 | 154360797, 154380719, 154382139, 154393680, 154396472 | *OPRM1* | Cross-ancestry (Less stringent), EA (Less Stringent), Cross-ancestry (Stringent), EA (Stringent), Cross-ancestry (Meta), EA (Meta) |
| 6 | 24394925 | *MRS2* | HA (Less stringent) |
| 8 | 143312933, 143316970 | *TSNARE1* | Cross-ancestry (Stringent), EA (Stringent), EA (Meta) |
| 9 | 127873473, 127959540, 127980426 | *SCAI, RABEPK* | Cross-ancestry (Less stringent), Cross-ancestry (Meta), EA (Meta) |
| 10 | 103414885 | *FBXW4* | Cross-ancestry (Less stringent), EA (Less Stringent), EA (Meta) |
| 10 | 110504365 | [ ] | Cross-ancestry (Less stringent) |
| 11 | 112869404 | *NCAM1* | Cross-ancestry (Less stringent) |
| 15 | 91410009, 91406146, 91426560 | *FURIN* | Cross-ancestry (Less stringent), EA (Less Stringent), Cross-ancestry (Stringent), EA (Stringent), Cross-ancestry (Meta), EA (Meta) |
| 16 | 61631362 | *CDH8* | EA (Less stringent), EA (Stringent) |
| 19 | 18093588 | *KCNN1* | Cross-ancestry (Less stringent), AA (Less stringent), Cross-ancestry (Meta), AA (Meta) |
| 20 | 48540277, 48583726 | *RNF114* | Cross-ancestry (Less stringent), Cross-ancestry (Meta) |

### SNP Heritability and Genetic Correlations Across GWAS Datasets

The estimated SNP heritability (h^2^_SNP_) in MVP for the less stringent phenotype is similar in AAs (0.11 ± 0.03) and EAs (0.12 ± 0.01). Using the stringent OUD phenotype, the estimated h^2^_SNP_ was higher in AAs (0.20 ± 0.05) than EAs (0.15 ± 0.01). In contrast, the estimated h^2^_SNP_ was higher in EAs (0.11± 0.01) than AAs (0.08 ± 0.03) in the ancestry-specific meta-analyses across datasets (Supplementary Table 9). Variation in these estimates appears to be mainly driven by changes in effective sample size, as estimates using actual sample size show little variation (Supplementary Table 9). The within-ancestry r_g_ between datasets is high, ranging from 0.7 (± 0.3) between the less stringent OUD MVP and Partners HealthCare Biobank datasets in EAs, to 1.2 (± 0.2) between the less stringent OUD MVP and the previous OUD MVP GWAS^13^ in EAs (which used the same diagnosis definition as the present stringent analysis) (Supplementary Table 10).

Considering the similarity in h^2^_SNP_ between the different OUD GWAS and the greater number of loci captured by the less stringent diagnosis in MVP, all subsequent downstream analyses are based on the less stringent OUD GWAS within the MVP sample.

### Partitioning Heritability Enrichment

We performed partitioning heritability enrichment analyses in LDSC^27^ and examined heritability enrichment for gene expression using GTEx data^40^. The most significantly enriched cell type group was the central nervous system (CNS; *p* = 3.34 × 10^−3^, Figure 2A, Supplementary Table 11). We observed significant enrichment for OUD in brain tissues only, including the anterior cingulate cortex (*p* < 4.71 × 10^−6^), limbic system (*p* < 3.25 × 10^−5^), prefrontal cortex (*p* < 5.73 × 10^−5^), cerebral cortex (*p* < 9.81 × 10^−5^), cortex (*p* < 1.11 × 10^−4^), hypothalamus (*p* < 1.23 × 10^−4^), amygdala (*p* < 1.41 × 10^−4^), and hippocampus (*p* < 2.04 × 10^−4^) (Figure 2B, Supplementary Table 12). There were no significant enrichments for epigenetic annotations after correction for multiple testing (Supplementary Table 13).

**Figure 2:**
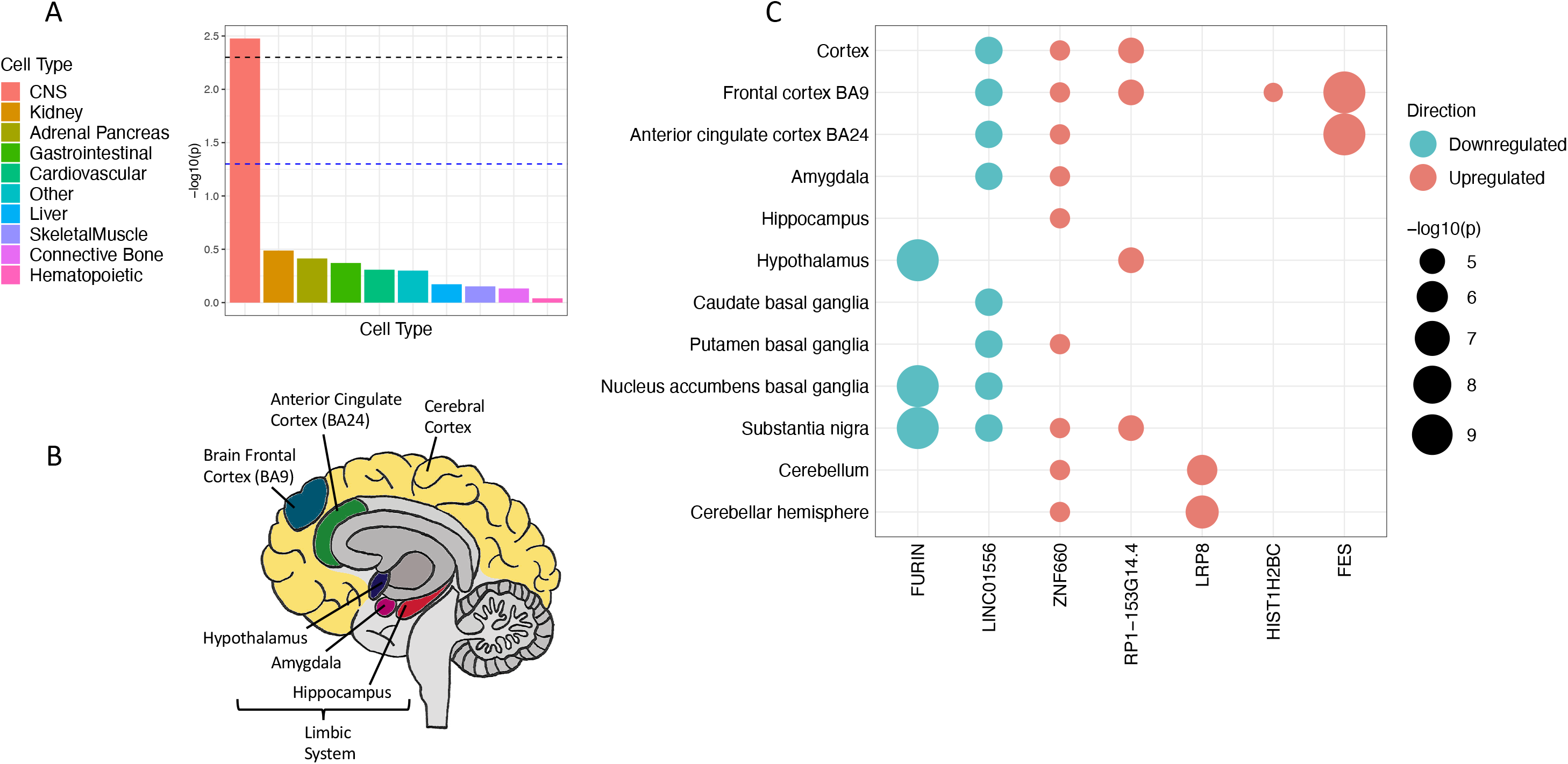
Enrichment of OUD in the brain. A. Partitioning heritability enrichment analyses using LDSC show enrichment for OUD in the central nervous system. B. Heritability enrichment for gene expression using GTEx data show enrichment for OUD in brain regions previously associated with addiction. C. Predicted gene expression using S-PrediXcan identify genes with differential expression in brain regions.

### Transcriptome-wide Analysis

We used S-PrediXcan^32^ to predict the effect of genetic variation on gene expression. Significant within-tissue gene expression regulation was identified for 43 tissues, including brain, adipose, gastrointestinal, thyroid, and liver (Supplementary Figure 2, Supplementary Tables 14 and 15). Significant associations with expression in brain tissues were detected for *FURIN*, *FES*, *LRP8*, *LINC01556*, *ZNF660* and *RP1-153G14.4* (Figure 2C). Some of these genes (*FURIN*, *LINC01556*, *ZNF660*, and *RP1-153G14.4*) were also expressed in non-brain tissues, such as adipose, gastrointestinal, and thyroid tissues (Supplementary Figure 2), suggesting that OUD-related genetic variation may exert significant transcriptomic changes in the periphery as well as the CNS.

Considering the sharing of eQTLs across multiple tissues, we tested the joint effects of variation in gene expression across tissues using S-MultiXcan^34^. Significant transcriptomic effects for OUD were detected in 8 genes, 5 of which overlapped with genes detected by S-PrediXcan (*FURIN*, *FES*, *RP1-153G14.4*, *LRP8*, and *RABEPK*) and 3 which were novel (*ZNF391*, *ZKSCAN4*, and *MAGOH*) (Supplementary Table 16).

### Gene Set, Functional Enrichment, and Drug Repurposing Analyses

Gene-based analyses identified one GWS gene in AAs (*CHRM2*, p = 9.52 × 10^−7^) and three GWS genes in EAs (*OPRM1*, p = 2.17 × 10^−7^; *FTO*, p = 9.52 × 10^−7^; *DRD2*, p = 1.67 × 10^−6^) (Supplementary Figure 3), but none in HAs. Following Bonferroni correction, no biological processes or pathways were significantly enriched, although nominal associations in EAs highlighted pathways of potential relevance, including “dopamine receptors” (p = 1.87 × 10^−5^) and “regulation of adenylate cyclase activating G-protein coupled receptor signaling pathway” (*p* = 4.39 × 10^−5^) (Supplementary Table 17).

Genes identified in the variant-level, gene-based, or transcriptome (brain region) analyses (N=24) are summarized in Supplementary Table 18. Examination of these genes for drug-gene interactions via the Drug Gene Interaction database identified 761 interactions between 8 genes (*CHRM2, DRD2, FES, FURIN, KCNN1, NCAM1, OPRM1, PRL*) and 340 unique medications (Supplementary Table 19, Supplementary Figure 4). *OPRM1* had 193 interactions, mainly with classes of analgesics, anesthetics, and drugs for constipation. *DRD2* had 376 interactions, the majority of which were with psycholeptics.

### Genetic Correlations

We estimated pairwise *r*_g_ with OUD for 40 published phenotypes using LDSC^25^. OUD showed significant *r*_g_ with 21 traits. As expected, the strongest positive correlations were with substance use traits (e.g., problematic alcohol use, cannabis use disorder, ever smoked regularly) and psychiatric disorders (e.g., bipolar disorder, major depressive disorder) and the strongest negative correlation was with educational attainment (Figure 3A, Supplementary Table 20). We also assessed *r*_g_ with OUD for 1,270 complex traits from the UKBB using CTG-VL^36^. After multiple testing correction (*p* < 3.94 × 10^−5^), 106 traits demonstrated significant genetic association with OUD (Supplementary Figure 5, Supplementary Table 21). These included positive correlations with substance use related traits (e.g. current smoking, ever addicted to any substance or behavior), psychiatric traits (e.g. anxiety treatment, self-reported depression) and pain-related traits (e.g. low back pain, multisite chronic pain), and negative correlations with having secondary education qualifications and the presence of social support. Thus, overall, we found that increased risk of OUD is genetically correlated with increased liability for use of substances, psychiatric disorders, and experiencing pain, and lower likelihood of educational attainment and social support.

**Figure 3:**
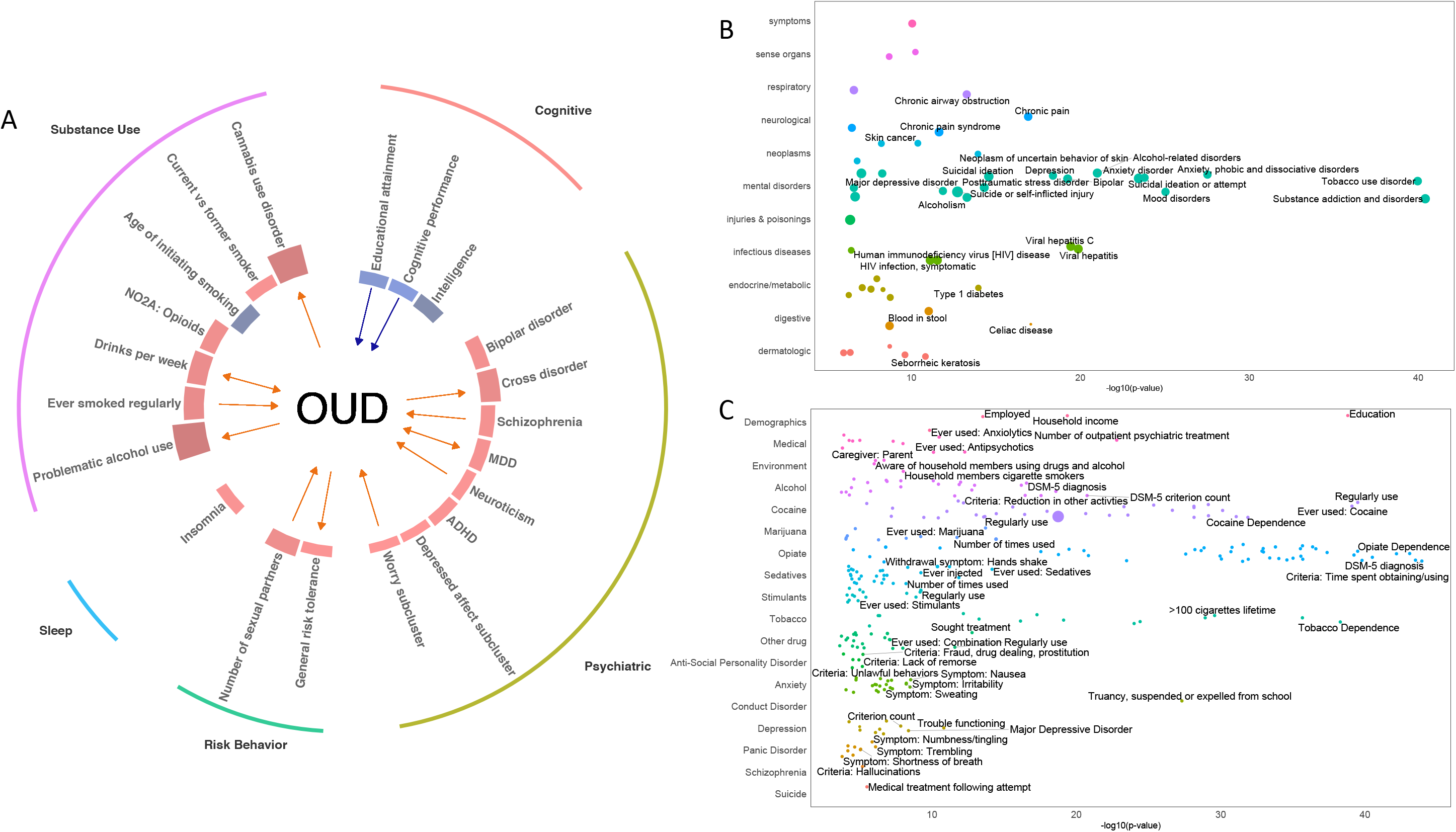
Phenotypic spectrum associated with OUD. A. Genetic correlation analyses show multiple traits significantly genetically correlated with OUD (red bar – positively correlated, blue bar – negatively correlated). Mendelian randomization analyses identify causal associations between OUD and other traits (arrows, red – positive causal association, blue – negative causal association). B and C. PheWAS results in BioVU (B) and Yale-Penn (C) datasets. All phenotypes significant at FDR p<0.05 are plotted. In B, all phenotypes which pass Bonferroni correction are labelled. For readability, in C, only the top 3 traits within each group which pass Bonferroni correction are labelled. Circle size denotes effect size.

### Mendelian Randomization

Using MR, we tested for bidirectional causal effects between OUD and the 21 traits identified as significantly genetically correlated with OUD (Figure 3A, Supplementary Figure 6). There was a causal effect of OUD on 6 traits: problematic alcohol use, drinks per week, cannabis use disorder, general risk tolerance, MDD, and cross disorder. Among the 21 traits, 9 had a causal effect on OUD, of which 2 showed a negative causal effect on OUD (cognitive performance and educational level) and 7 a positive causal effect on OUD (in descending order of magnitude: drinks per week, worry subcluster, neuroticism, the number of sexual partners, major depressive disorder, cigarettes per day and schizophrenia).

### Polygenic Risk Scores and Phenome-wide Association Studies

PRS were calculated in 2 independent datasets to identify phenotypic associations of genetic liability for OUD. In the Yale-Penn sample, PRS were calculated for 4,922 African ancestry and 5,709 European ancestry individuals. No significant associations were identified for AAs (Supplementary Figure 7, Supplementary Table 22). In EAs, PheWAS identified 41 phenotypes in the opiate domain and 71 phenotypes in other phenotypic domains that were significantly associated with OUD PRS (Figure 3C, Supplementary Table 23). The most significantly associated phenotypes were “time spent obtaining/using opioids” and “opioid use disorder”. In BioVU, PRS were calculated for 12,384 AAs and 66,903 EAs. No significant associations were found for OUD PRS in AAs (Supplementary Figure 8, Supplementary Table 24). In EAs, the OUD PRS was associated with 27 phenotypes, including “substance addiction and disorders” and “mood disorders” (Figure 3B, Supplementary Table 25).

### Genomic Structural Equation Modeling

We conducted genomic structural equation modeling (gSEM) to evaluate how OUD relates to the three other substance use traits and the seven psychiatric disorders identified as the most significantly associated with OUD in genetic correlation analyses. Exploratory factor analysis (EFA) involving all 11 traits supported a 4-factor model with cumulative variance of 0.639. We retained paths with a loading factor ≥0.2 and conducted confirmatory factor analysis (CFA). In this analysis, the 4-factor model fit the data well (comparative fit index (CFI) = 0.948, Akaike information criterion = 340.840, χ^2^ = 276.840, degrees of freedom = 34, standard root mean root square error (SRMR) = 0.073). The 4 substance use traits all loaded on Factor 1, with a major contribution from OUD (loading = 0.84 ± 0.05) and problematic alcohol use (loading = 0.91 ± 0.3), and lower contributions from cannabis use disorder (loading = 0.58 ± 0.06) and ever smoked regularly (loading = 0.40 ± 0.03). Cannabis use disorder (loading = 0.37 ± 0.06) and ever smoked regularly (loading = 0.42 ± 0.03), together with other psychiatric disorders, also loaded on Factor 3. Major psychiatric disorders, including bipolar disorder (loading = 0.86 ± 0.04), schizophrenia (loading = 0.76 ± 0.03), and MDD (loading = 0.43 ± 0.03) loaded on Factor 2. Tourette’s syndrome (loading = 0.33 ± 0.07) and obsessive-compulsive disorder (loading = 1.03 ± 0.21) loaded on Factor 4 (Figure 4, Supplementary Table 26).

**Figure 4:**
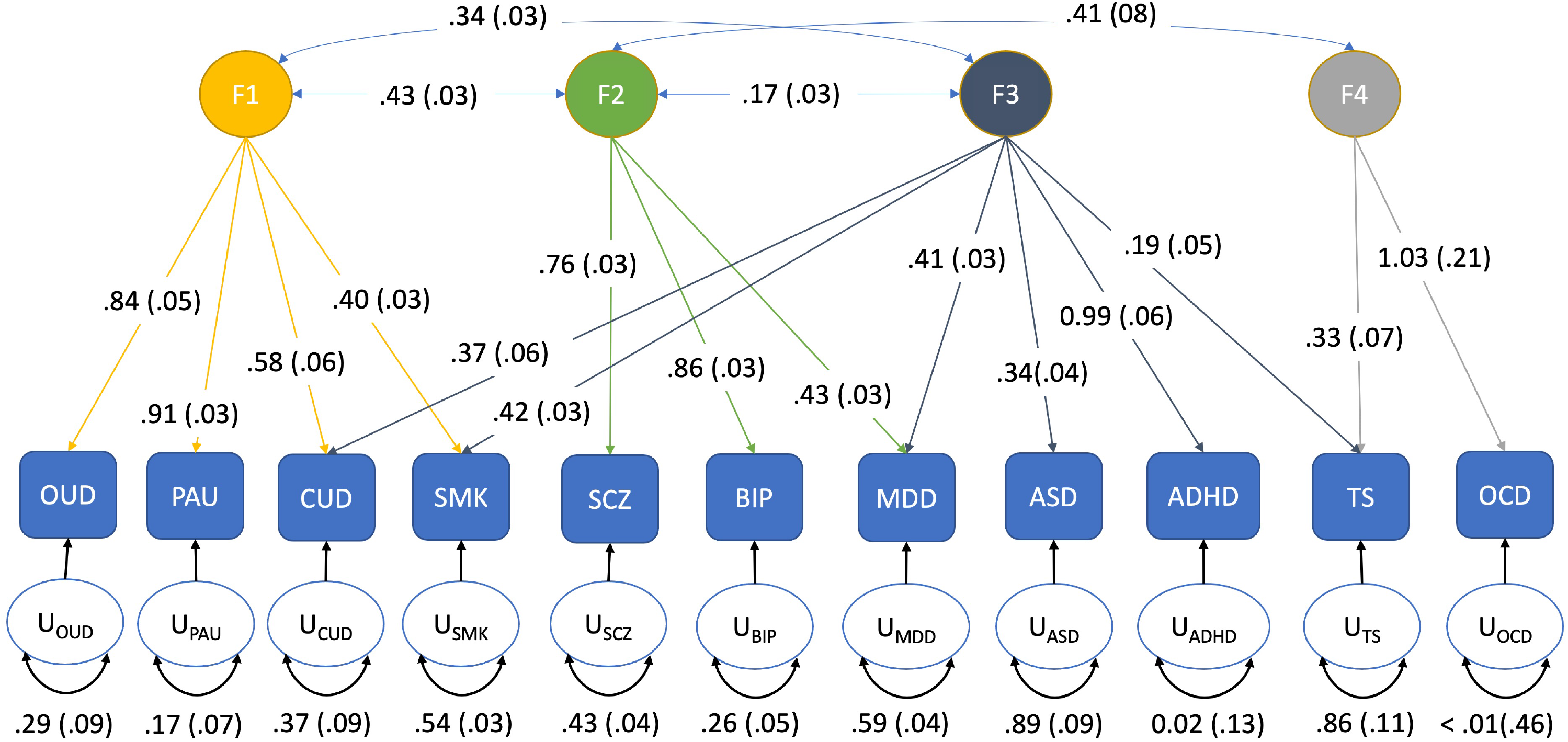
Genomic SEM analysis of OUD with other substance use traits (OUD: opioid use disorder; PAU: problematic alcohol use; CUD: cannabis use disorder; SMK: ever smoked regularly) and psychiatric disorders (SCZ: schizophrenia; BIP: bipolar disorder; MDD: major depressive disorder; ASD: autism spectrum disorder; ADHD: attention deficit hyperactivity disorder; TS: Tourette’s syndrome; OCD: obsessive compulsive disorder). Four factors were identified. Factor loadings for each trait are depicted by arrows between the trait and the factor. Correlation between factors is indicated by arrows between the factors. Residual variance for each trait is indicated by the U-circles. Standard errors are depicted in parentheses.

## Discussion

This study, the largest single-sample GWAS of OUD to date, identified 14 loci associated with the disorder, 12 of which are novel findings. Three of these loci were significant in ancestry-specific analyses only, demonstrating that inclusion of diverse ancestral samples in genetic studies of OUD permits the identification of novel genetic variants. Post-GWAS analyses in EAs revealed enrichment for OUD in the CNS, particularly the brain, and an extensive phenotypic spectrum associated with genetic liability for OUD.

Because the effect sizes of common variants contributing to highly polygenic phenotypes such as OUD are small, large sample sizes are required to identify GWS loci. The largest OUD GWAS prior to the current study greatly increased the effective sample size (N_effective_ = 88,115) by meta-analyzing the results of studies that used a range of case and control definitions^16^. Here, we performed GWAS using the stringent definition of OUD used by Zhou et al.^13^ (N_cases_ = 23,459, N_effective_ = 88,569) and a less stringent definition requiring the presence of only 1 ICD-9/10 diagnostic code for opioid abuse or dependence (N_cases_ = 31,473, N_effective_ = 116,590). Although the less stringent definition lowers the specificity of the case phenotyping (i.e., individuals are more likely to be mislabeled as having OUD), it increases the number of cases by more than 8,000 and reveals 8 more GWS variants than the stringent definition. These results support prior conclusions that the potential variability introduced by broadening phenotypic definitions in genetic studies of OUD is outweighed by substantial increases in sample size^16^; however owing to clinical differences between substance use disorders and other psychiatric disorders, this may not be generalizable. In contrast, our meta-analysis of the MVP data with other datasets reduced the number of GWS loci identified, potentially because the smaller additional datasets increased the variability in the effect size of variants, reducing associations overall.

The most significant locus, *OPRM1*, encodes the mu-opioid receptor, which binds morphine and other opioids and has been the focus of many functional and candidate gene studies of opioid-related phenotypes^41–43^. In a previous GWAS comprised principally of subjects from MVP, OUD was significantly associated only with *OPRM1* in EAs^13^, with the lead SNP being the non-synonymous, exon 1 variant rs1799971 (A118G). In neither that study, nor the present study, was the SNP associated with OUD in AAs, presumably because the minor (G) allele frequency in this population group is considerably lower than in EAs^44^. Even so, it is difficult to explain why meta-analysis with AAs does not increase the statistical strength of the association of OUD with this variant if it is truly the lead functional variant, even if based on introgressed EA alleles alone. A recent meta-analysis of opioid addiction in European-ancestry individuals, which included data from the previous MVP OUD GWAS^13^ and 4 other cohorts also identified *OPRM1* as a risk gene, although the lead SNP at that locus was rs9478500 in intron 1^16^.

We identified a second peak in *OPRM1*, with the lead SNP rs3778151, a variant in intron 1 that is in high LD with rs9478500 (r^2^ = 0.56-0.90)^45^, the variant associated with opioid addiction in the recent meta-analysis^16^. Although LD clumping in our sample showed these two loci to be independent, when we conditioned the analysis on rs1799971, rs3778151 was no longer significant, suggesting that the variants are structurally independent, but functionally redundant. A prior study in EA alcohol- or drug-dependent cases and controls also identified two independent LD blocks in *OPRM1*^46^. The findings were interpreted as a partial explanation of the inconsistent findings for *OPRM1*, particularly rs1799971, in candidate gene studies of alcohol and drug use disorders^46^. Our findings suggest that although LD clumping may differentiate the two *OPRM1* loci, a more definitive test of independence – namely, a conditional analysis – fails to show their independence, consistent with the presence of a single locus at *OPRM1*.

Our cross-ancestry analysis showed an association between a SNP in *FURIN* and OUD, with transcriptome-wide analyses showing significant downregulation of gene-expression for *FURIN* in brain-related tissues. These findings support the reported associations of OUD with *FURIN* both in gene-based analyses^14, 16^ and in a variant-level meta-analysis of OUD^14^. *FURIN* is associated with schizophrenia^47^ and harbors a schizophrenia-associated cis-eQTL^48^. Although *FURIN* encodes a protease that cleaves some endogenous opioids, notably proenkephalin^49^, the enzyme has not previously been linked to the effects of exogenous opioids or mu-opioid receptor signaling. Given these findings, further research on the mechanism underlying the gene’s effects on risk of OUD is warranted.

Our analysis also identified 12 novel GWS loci. Two of these, which are in *RABEPK* and *NCAM1*, were GWS in a multi-trait analysis using MTAG of OUD with cannabis use disorder and alcohol use disorder^14^. Here, we show associations directly with OUD. *RABEPK* is adjacent to *PPP6C*, a gene previously implicated in a gene-level analysis of OUD^16^ that has also been linked to reward-related phenotypes like obesity and smoking^50, 51^. Whereas SNPs within *RABEPK* contributed to the *PPP6C* signal in a previous study^45^, our lead SNP could be tagging functional variants in *PPP6C*. Alternatively, our study included multiple ancestral groups, and 50% more OUD cases, which could have affected the location of the GWAS peak and enabled us to localize the peak more accurately than in ^45^. *NCAM1* and *KCNN1* have been implicated in the neuropharmacology of opioid-related phenotypes. The mouse homolog of *KCNN1* is differentially expressed in the nucleus accumbens following chronic morphine exposure^52^. The gene is also downregulated in the rodent prelimbic cortex after exposure to cues associated with morphine withdrawal^53^, suggesting a connection to learning and memory. *NCAM1* also appears to be involved in the response to morphine exposure. Tolerance in rodents due to repeated morphine injection can be prevented by treatment with an antisense oligodeoxynucleotide that targets *Ncam1*^54^. *NCAM1* variants have also been significantly associated with alcohol dependence^55^, drug dependence^56^ , smoking initiation^51^, cannabis use^57^, and alcohol consumption^51^.

Several other loci identified in our cross-ancestry meta-analysis contain GWAS hits for other traits, suggesting the possibility of widespread pleiotropy of loci associated with OUD. *CDKAL1* and *BTNL2* have been associated with metabolic traits such as type 2 diabetes, body mass index^58^ and obesity-related phenotypes^50^. *FBXW4* and *CDH8* have prior associations with cognitive traits such as educational attainment and mathematical ability^59^, and *TSNARE1* has a prior association with schizophrenia^60^.

Partitioning heritability enrichment analyses showed that CNS cells were the only significantly enriched group. We found significant enrichment for OUD in brain tissues only, including the anterior cingulate cortex, limbic system, prefrontal cortex, hypothalamus, amygdala, and hippocampus, regions previously associated with the underlying neurobiology of the disorder^61^. These findings underscore the neural basis of OUD and reinforce the conceptualization of substance use disorders, which are often chronic and relapsing, as brain diseases. This notion was novel when proposed nearly 25 years ago^62^ and although today it is a view widely held by neuroscientists and clinicians, it is not universally understood by politicians or the general public. Improving our understanding of the biological basis of OUD could promote a science-based response to the opioid epidemic.

Consistent with prior findings, OUD showed strong genetic correlations with multiple substance use disorders, psychiatric disorders, cognitive traits, and risk behavior^13, 15, 16, 63^. Mendelian randomization analyses demonstrated causal effects of OUD on problematic alcohol use and cannabis use disorder, and a bidirectional causal effect of drinks per week. Given the high frequency with which substance use disorders co-occur, and the causal associations shown here, treatment efforts that aim broadly to reduce substance use are recommended: reducing use of substances comorbid with OUD would, according to this model, also reduce use of opioids. Genetic liability for psychiatric traits, including neuroticism and schizophrenia, was also causally associated with OUD, with a bidirectional causal effect of MDD on OUD. Our findings, along with those of others^13^, suggest that OUD has a common biological pathway with schizophrenia and MDD. Despite the significant genetic correlations and causal associations between OUD and psychiatric disorders, genomic structural equation modelling indicated a common genetic factor representing broad genetic liability for substance use disorders that is distinct from those underlying the psychiatric disorders. The factor structure among psychiatric disorders seen here is consistent with previous findings^64^ and shows that cannabis use disorder and smoking, unlike OUD, load onto both the substance use disorder factor and the factor underlying MDD, ADHD, autism spectrum disorder, and Tourette’s syndrome.

PheWAS of the genetic liability for OUD in the Yale-Penn sample, which was ascertained for substance use disorders, reproduced the broad association with other substance use. In a clinical dataset using EHR data, the genetic liability for OUD was associated with multiple traits in every phenotypic domain tested, demonstrating the widespread effects of OUD liability on bodily systems. Some of the associations may be due to phenotypic correlation. For instance, associations were found with viral hepatitis and human immunodeficiency virus (HIV), potential proxies of injection drug use, and with chronic pain and back pain, potential proxies for the use of analgesic medications. Negative associations with obesity, type 1 diabetes, and skin cancer may reflect under reporting or diagnosis in individuals with OUD, or they could also reflect true biological relationships.

Limitations to the present study should be noted. Although it includes AA, EA, and HA individuals, participants of European ancestry comprise more than 60% of the total sample, which in large part drove the results of the cross-ancestry analysis. This disparity in sample size is also reflected in analyses of the individual ancestral groups, in which the smaller AA and HA groups provided less statistical power and yielded fewer significant loci. The lower power of the AA GWAS is also reflected in the lack of associations in PRS analysis in AAs. Future GWAS of OUD should focus on expanding sample sizes for non-European-ancestral populations to capture loci that are relevant to specific population groups. The sample for this study is >90% male, reflecting the sex distribution of Veterans in the United States. Risk variants relevant only to women may thus have been overlooked due to the lower statistical power. Because the MVP dataset lacks information on the initiation of opioid use among patients diagnosed with OUD, we could not differentiate patients who developed the disorder only after being prescribed opioid analgesics from those whose first opioid use involved recreational use of analgesics or heroin. Differences in the initiation of opioid use could reflect different genetic risk factors contributing to non-overlapping intermediate phenotypes (e.g., pain threshold/susceptibility in analgesic use vs. risk taking in recreational use).

In summary, we have identified 14 genetic loci associated with OUD, the majority novel. Many of the loci contain genes with prior associations with substance use or psychiatric disorders, suggesting widespread pleiotropy. The use of a less stringent definition of OUD allowed a 25% larger number of OUD cases than a stringent definition in the MVP sample. Downstream analyses validate this approach by demonstrating plausible enrichment of OUD in brain regions, genetic correlations with other substance use disorders and psychiatric disorders, and association between OUD PRS and opioid dependence in an independent sample. Our findings provide insight into the biological underpinnings of OUD, which could inform preventive, diagnostic, and therapeutic efforts and thereby help to address the opioid epidemic.

## Data Availability

The full summary-level association data will be made available through dbGaP upon publication

## Acknowledgments

This work was supported by Merit Review Awards from the U.S. Department of Veterans Affairs Biomedical Laboratory Research and Development Service (# I01 BX003341) and Clinical Science Research and Development Service (# I01 CX001734; the VISN 4 Mental Illness Research, Education, and Clinical Center; NIAAA grant K01 AA028292 (to RLK); and NIDA grant DA046345. The views expressed in this article are those of the authors and do not necessarily represent the position or policy of the Department of Veterans Affairs or the United States Government.

