## Supplementary Figures for "Cross-ancestry meta-analysis of opioid use disorder uncovers novel loci with predominant effects on brain"

Supplementary Figure 1: Overview of analyses

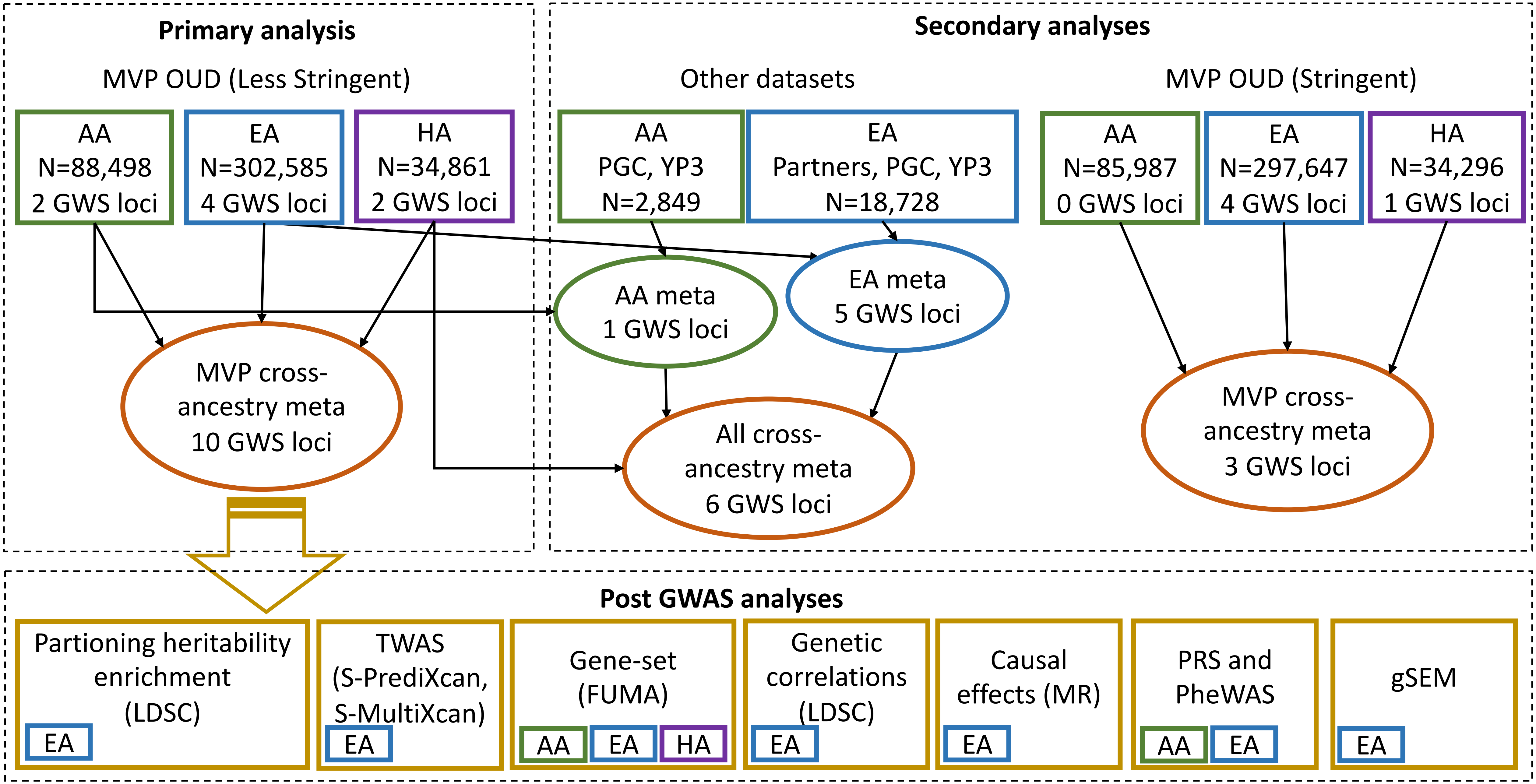


Summary of all analyses. Top left: Primary analysis for OUD (less stringent) in MVP dataset. Within ancestry GWAS for African American (AA), European American (EA) and Hispanic American (HA) followed by cross-ancestry meta-analysis. These results were used for all downstream analyses. Top right: Supplementary analyses. 1. Within ancestry meta-analysis was conducted for AA between MVP, PGC, YP3 and for EA between MVP, Partners, PGC, YP3, followed by cross-ancestry meta-analysis. 2. Within ancestry OUD (stringent) GWAS for African American (AA), European American (EA) and Hispanic American (HA) followed by cross-ancestry meta-analysis. Bottom: Post GWAS analyses were conducted in Aas, EAs and HAs as indicated.

Supplementary Figure 2: Within-tissue gene expression (S-PrediXcan)

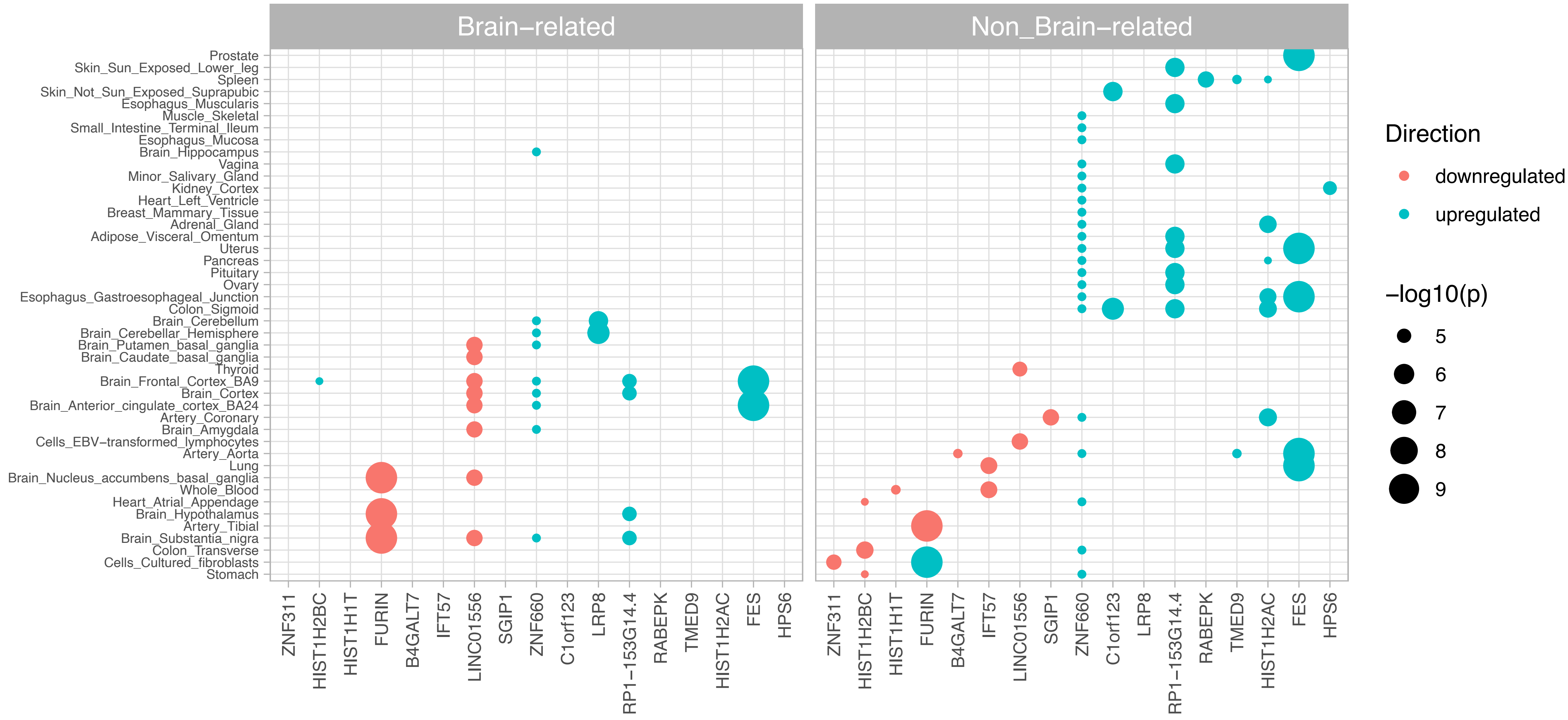


Within-tissue gene expression conducted using S-PrediXcan. Genes with predicted differential expression in brain-related tissues (left) and non-brain related tissues (right). Color of circle indicates downregulation (red) or upregulation (blue). Size of circle indicates –log10 p-value.

Supplementary Figure 3: Gene-based analyses

AA

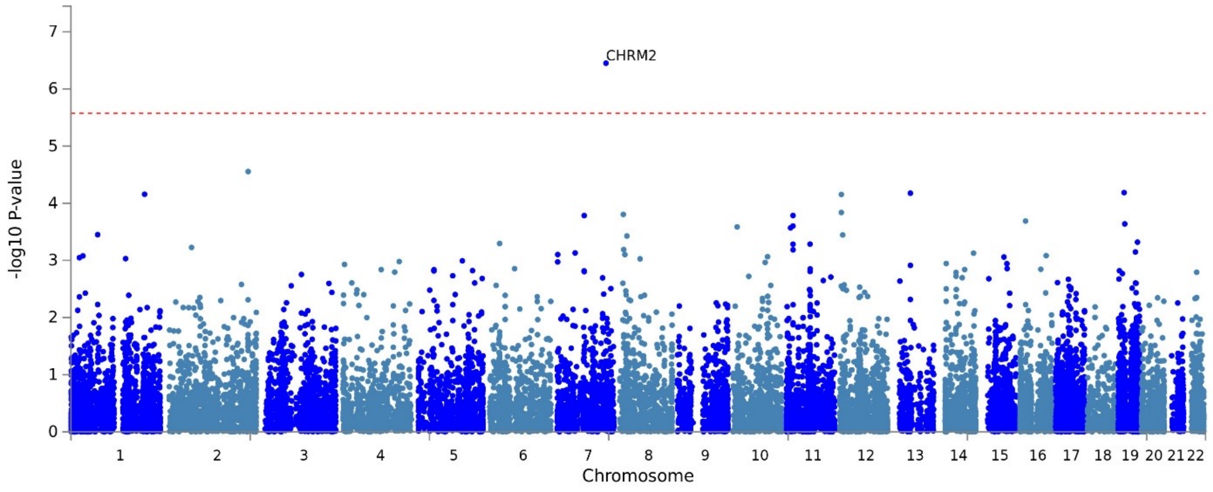


HA

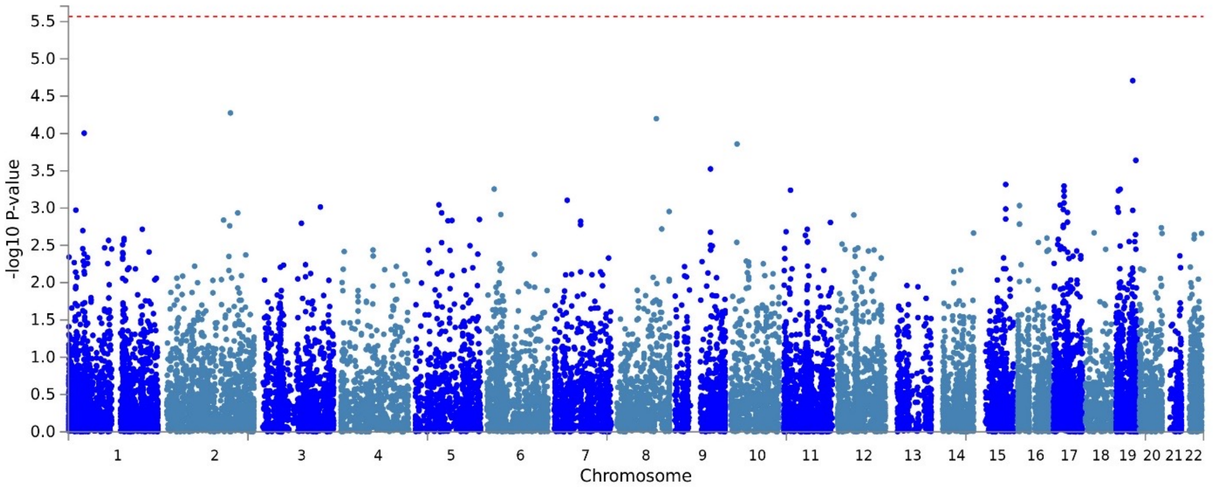


EA

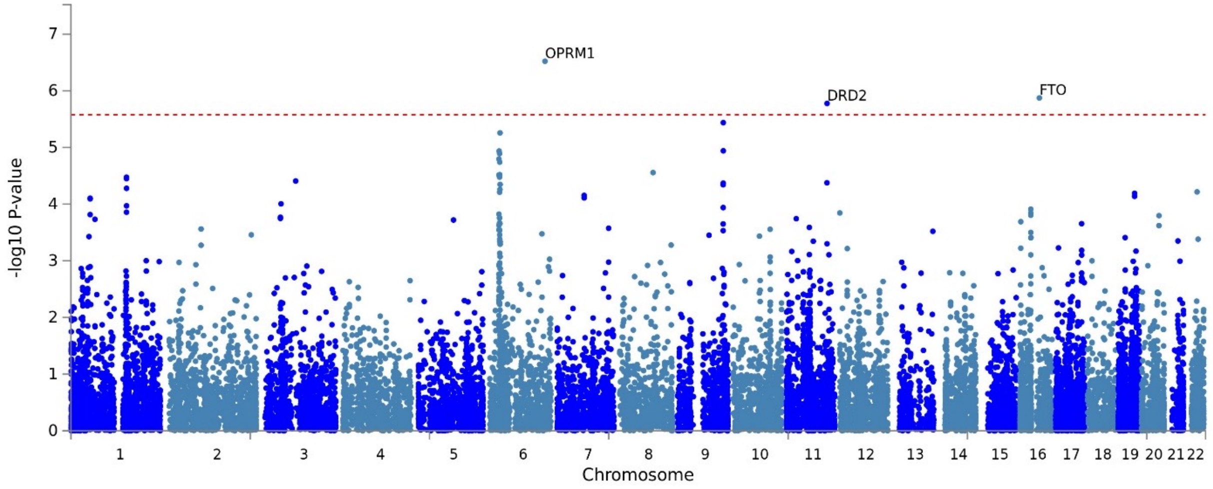


Gene-based Manhattan plots for AA, EA and HA. Gene-based association analyses were conducted using FUMA.

Supplementary Figure 4: Drug-gene interactions

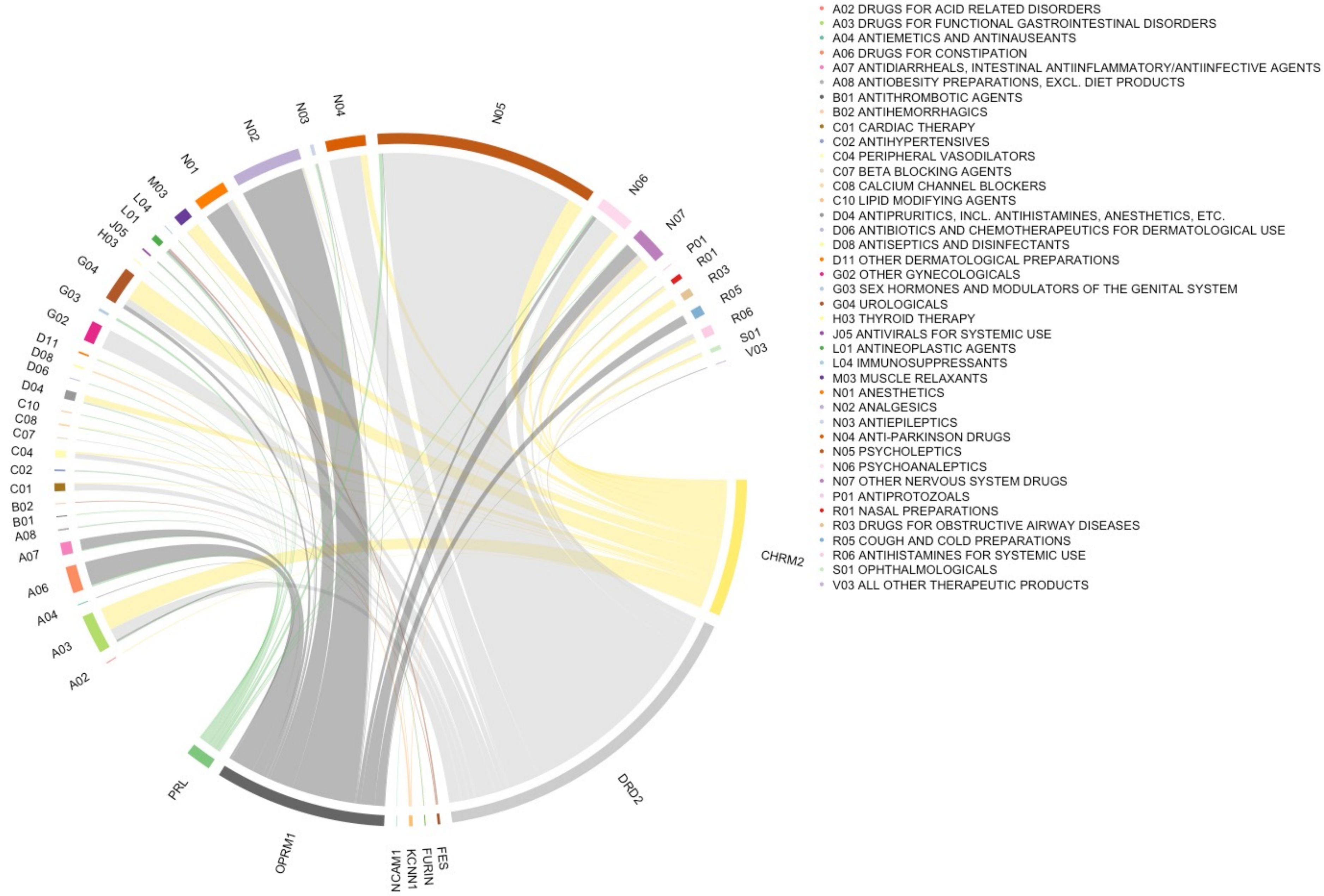


Drug-gene interactions for 8 genes associated with OUD. The width of the line between each gene and drug class indicates the number of interactions.

Supplementary Figure 5: Genetic correlation between OUD and UKBB complex traits

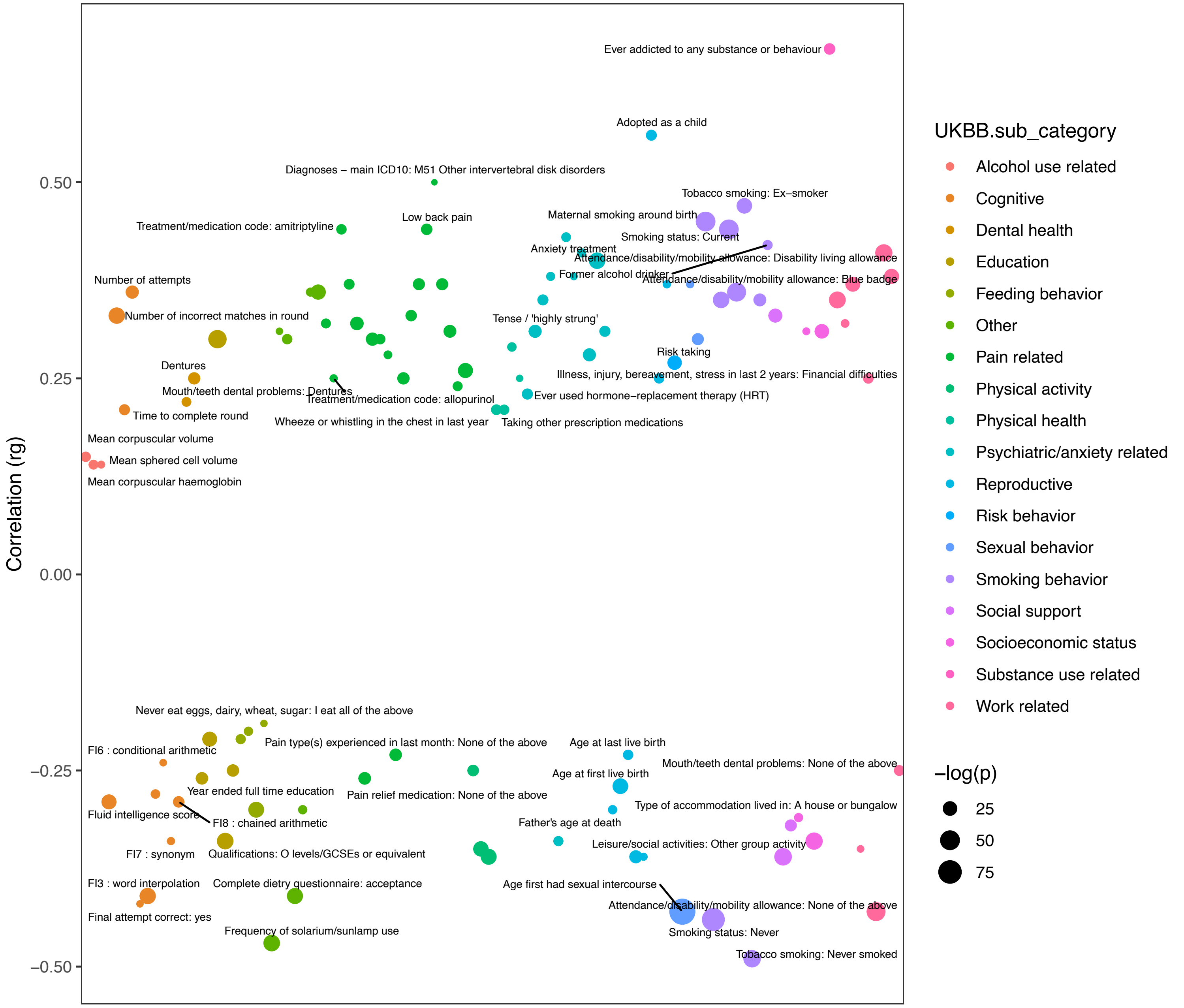


Genetic correlations for OUD in the UK Biobank dataset. All points passing Bonferroni correction are plotted. The color of the circle indicates the phenotypic category. The size of the circle indicates the –log10 p-value.

### Supplementary Figure 6: Mendelian randomization

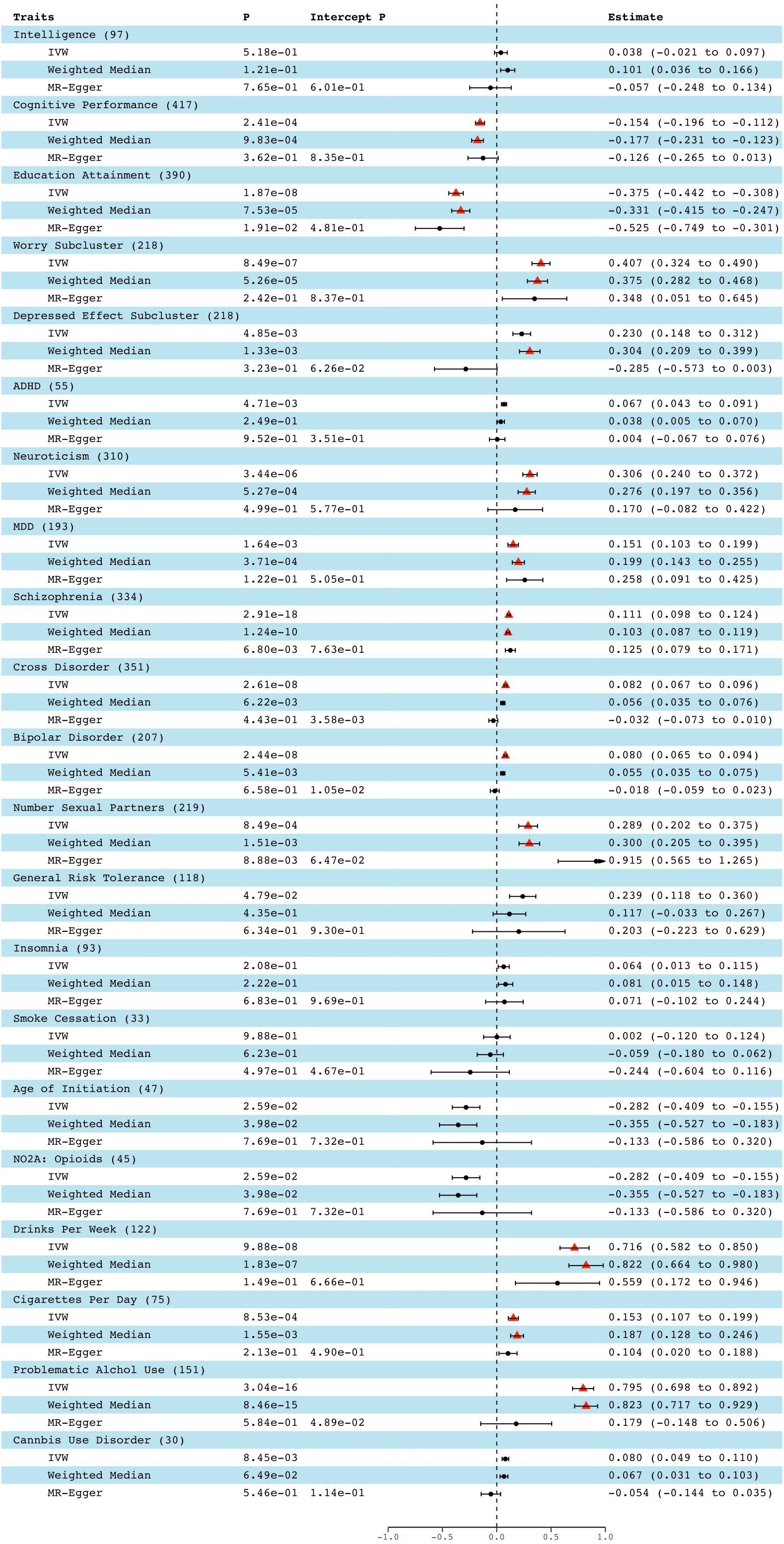


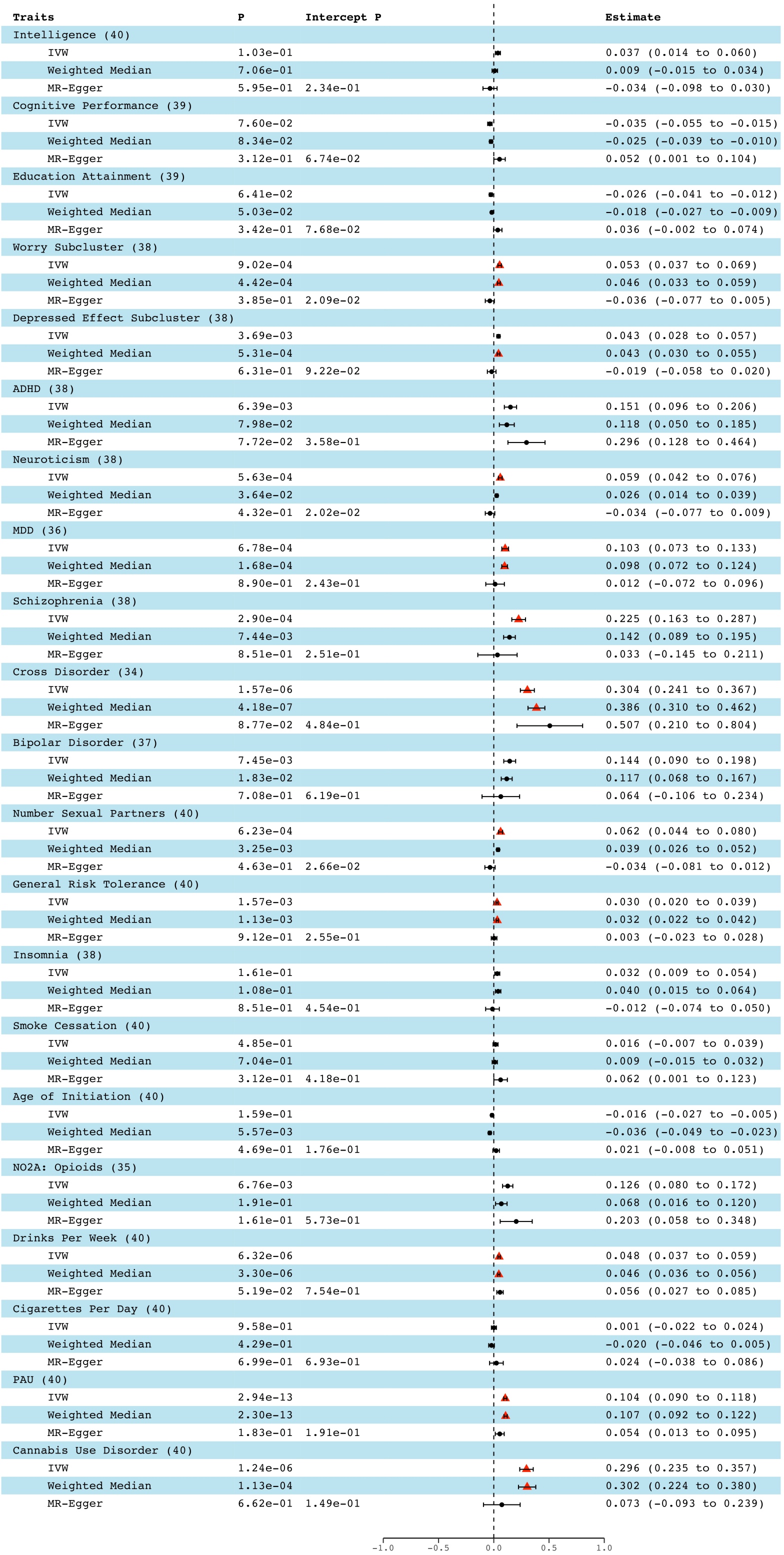


Causal association for traits genetically correlated with OUD. Left: Traits as the exposure, OUD as the outcome. Right: OUD as the exposure, traits as the outcome. Estimates and p-values for each MR analysis [Inverse variance weighted (IVW), Weighted median, and MR-Egger] are shown. Intercept p-value: MR-Egger horizontal pleiotropy test.

Supplementary Figure 7: PheWAS in Yale-Penn AA individuals

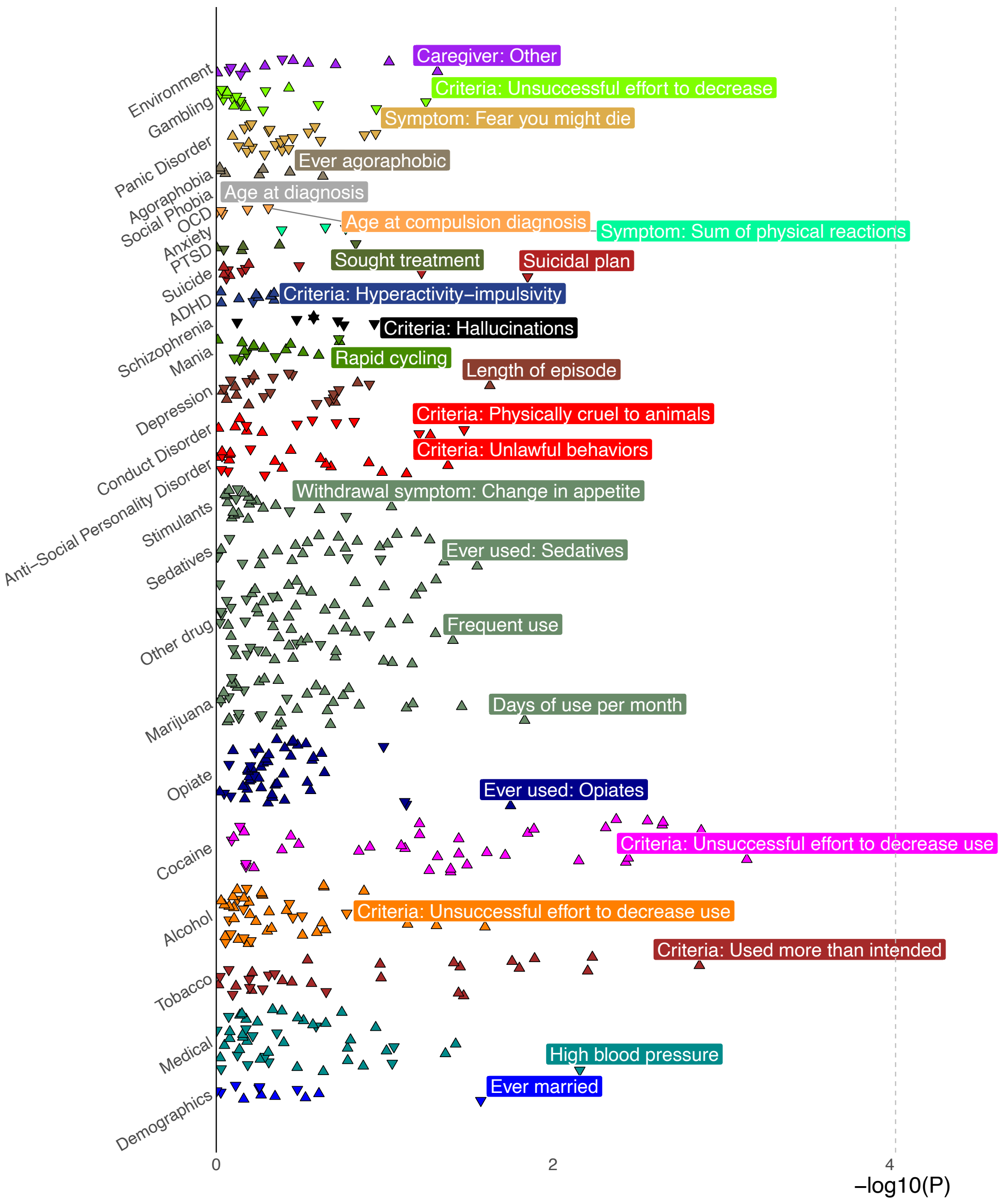


PheWAS plot for OUD PRS in AA individuals from Yale-Penn. No phenotypes pass Bonferroni correction.

Supplementary Figure 8: PheWAS in BioVU AA individuals

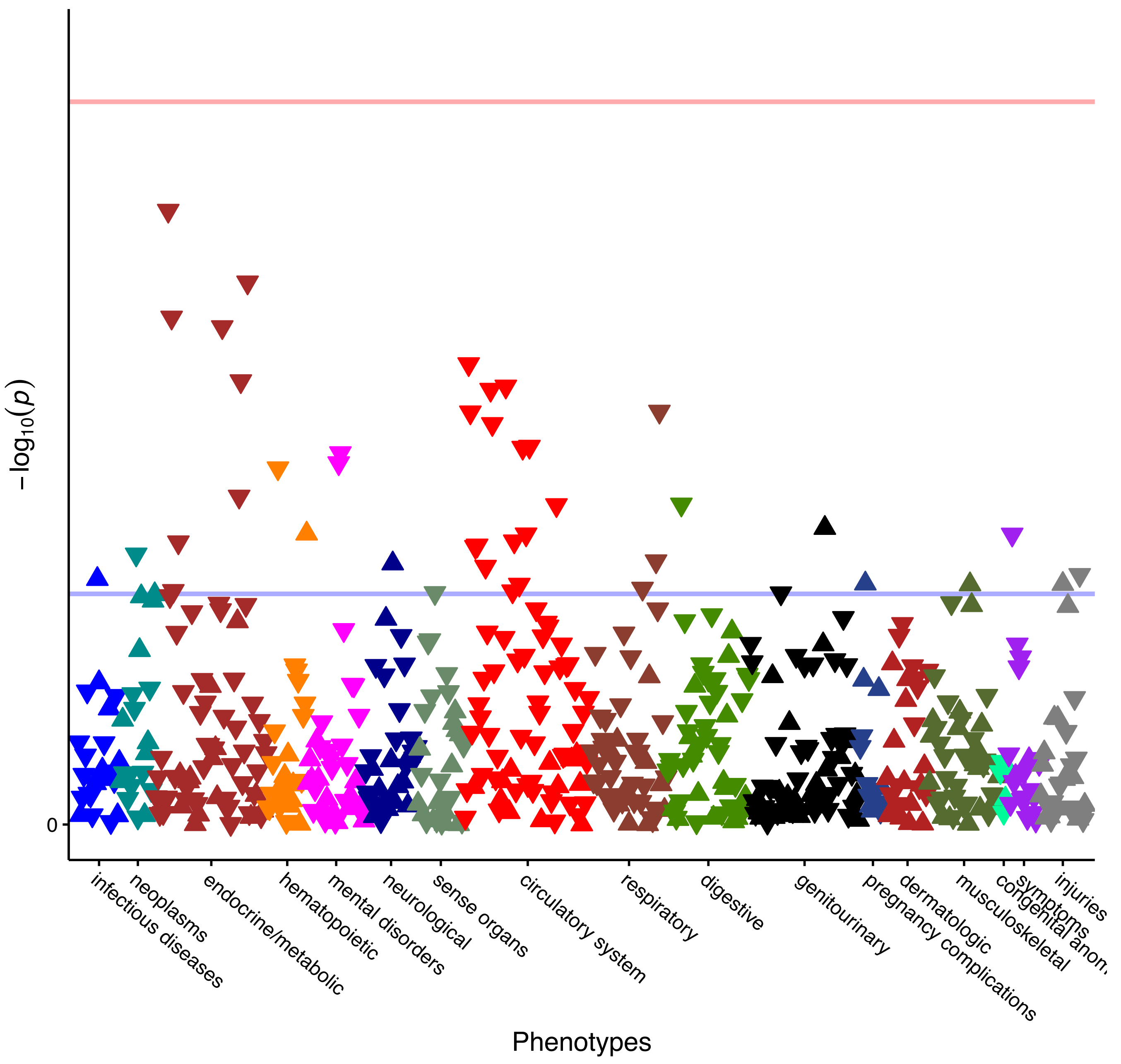


PheWAS plot for OUD PRS in AA individuals from BioVU. No phenotypes pass Bonferroni correction.
